# Documentation of compounded GLP-1 receptor agonists in a large primary care dataset

**DOI:** 10.1101/2025.05.12.25327436

**Authors:** Nathaniel Hendrix, Esther E. Velásquez, Harry Pham, Andrew Bazemore

## Abstract

Many Americans received compounded semaglutide and tirzepatide when these drugs were in shortage, but there is little understanding of how and how often they were used, limiting our ability to ensure safety, coordination, and appropriate regulation. We analyzed retrospective cohort data from the American Family Cohort from January 2021 to December 2024. We identified patients with documented semaglutide or tirzepatide use, categorizing them by brand and/or compounded drug exposure. Brand prescriptions were extracted from structured data; compounded drug use was identified from clinical notes. Among 153,075 included patients (64.0% female, mean age 55.0 years), 8.3% used compounded formulations. Compounded drug users had longer therapy durations versus brand-only users. Documentation rates of compounded semaglutide and tirzepatide use in primary care appear lower than survey-reported usage, suggesting that many patients received these medications outside coordinated care. Improved data interoperability, regulatory oversight, and clinical guidance are needed.

## Introduction

In 2022, two popular glucagon-like protein-1 receptor agonists (GLP-1 RAs), semaglutide and tirzepatide, were declared by the US Food & Drug Administration (FDA) to be in shortage. Compounding pharmacies satisfied much of the consumer demand that was left unmet.(1) Concerns quickly emerged around safety issues from some services offering compounded GLP-1 RAs, including misleading advertising and inadequate patient education.(2) In response to these problems, the American Diabetes Association published a statement recommending against the use of non-FDA-approved compounded GLP-1 RAs.(3)

The official shortages for tirzepatide and semaglutide ended in late 2024 and early 2025, respectively, bringing questions about the appropriateness of compounding laws to the fore. Despite the public health importance of compounded GLP-1 RAs over the three-year shortage, the decentralized nature of their distribution has also left many open questions about how they were used and how often. A survey found that, among the 12% of Americans who report having used GLP-1 RAs, 23% received prescriptions through aesthetic services or telehealth startups,(4) almost all of whom offer compounded instead of branded GLP-1 RAs.

In this study, we used a large, nationwide primary care database of extracted electronic health records to assess how often compounded GLP-1 RAs are documented and to characterize differences in users of compounded versus brand drugs. Our research questions in this descriptive cohort study were twofold. First, are primary care providers aware of their patients’ use of compounded GLP-1 RAs? And second, are users of brand GLP-1 RAs different from users of compounded versions?

## Methods

### Data source

Our data source was the American Family Cohort (AFC), a nationwide collection of electronic health records from primary care practices maintained by the American Board of Family Medicine in collaboration with Stanford University.(5) It contains records of over 5,000 providers’ encounters with nearly 8 million unique patients in 1,331 clinics. The analytic dataset was finalized March 12, 2025, though reporting lags mean that data was missing from some practices in the final months of 2024.

### Participants and exposures

Included patients received either semaglutide or tirzepatide between January 1, 2021 and December 31, 2024. While semaglutide initially entered the US market in 2017, the publication in 2021 of several high-profile clinical weight loss trials substantially increased demand.(6) Tirzepatide entered the US market in 2022, but already had published evidence of its weight loss impacts when it began sales.(7)

Brand drug prescriptions were available in structured data, but we had to consult clinical notes for mentions of compounded GLP-1 RAs. We iteratively developed a keyword-based search strategy aimed at achieving high positive predictive value. Our final, case-insensitive search strategy was as follows, where “%” indicates wildcard characters: [(“%semaglutide%”, “%ozempic%”, “%wegovy%”, “%tirzepatide%”, “%mounjaro%”, OR “%zepbound%”) AND (“%compound%”) AND NOT (“% compound %” OR “% compounds %”)] OR (“%semaglutide/%” OR “%tirzepatide/%”). Specifically, we excluded the stand-alone terms “compound” and “compounds” because they tended to appear in reference to supplements and in patient education around drug interactions (e.g., “avoid herbal supplements containing compounds like…”). We also found that many providers referred to compounded GLP-1 RAs containing added ingredients using a forward slash without explicit mention of the compounded nature of these drugs (e.g., “semaglutide/B12 injection”).

We estimated the positive predictive value of this method by reporting the number of false positives in a manual review of 500 random notes captured by the search strategy.

### Variables

We captured certain patient- and area-level variables to better characterize the populations using branded and/or compounded GLP-1 RAs. Specifically, we included gender, race/ethnicity, age, documented diagnosis of type 2 diabetes, whether the patient’s home address is in a state in which Medicaid covers GLP-1 RAs for weight loss,(8) and Reproducible Area Deprivation Index(9) of their home address. When available, we also included two population measures of hemoglobin A1c at initial prescription: mean A1c and percent of individuals with an A1c over 6.5%, which is the threshold for a diagnosis of type 2 diabetes.

### Statistical Methods

We separately evaluated inter-group differences across two stratified populations. First, we assessed differences between brand-only users, compounded-only users, and switchers. Second, we assessed differences between subgroups of switchers: those who initially used brand drugs, those who initially used compounded drugs, and those who had simultaneous documentation of both sources. We used ANOVA to assess the significance of differences in continuous values and chi-square tests for differences in categorical values.

### Data Sharing Statement

Complete data are available from the American Family Cohort for approved research projects. See up-to-date access requirements at https://www.AmericanFamilyCohort.org.

## Results

We identified 153,075 patients who received a prescription for an included GLP-1 RA (Figure 1). Of these, 127,024 used semaglutide and 53,142 used tirzepatide (17.7% used both). Overall, 140,446 (91.7%) used only brand drugs, while 6543 (4.3%) used only compounded drugs. Among the 6086 (4.0%) who had documented use of both drug sources, switching was common (Figure 2), with most (3725, 61.2%) starting a brand drug before switching to compounded formulations. Up to 2.5% of patients with documented use of an included medication in a given month switched between drug sources.

**Figure 1.**
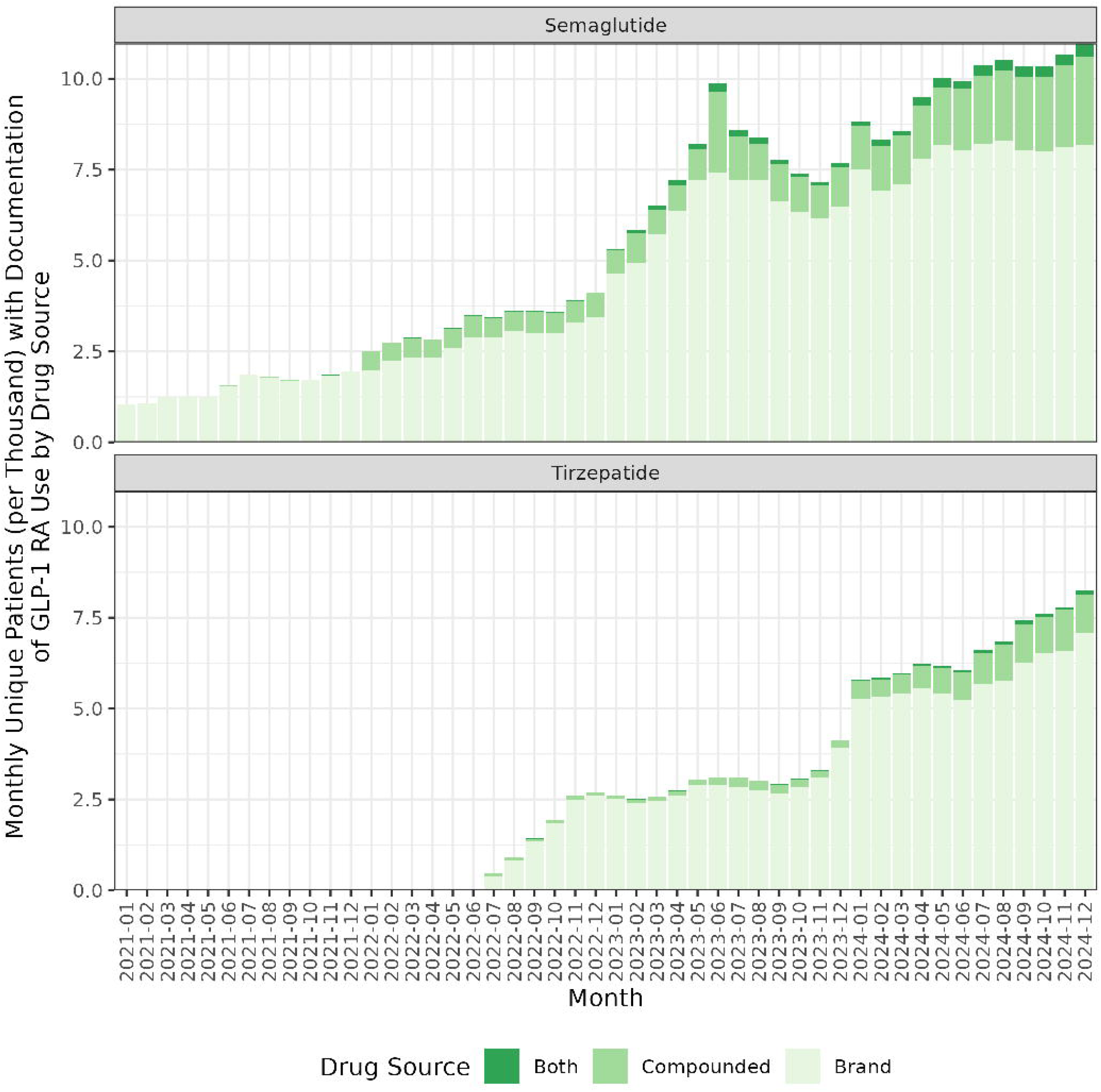

**Figure 2.**
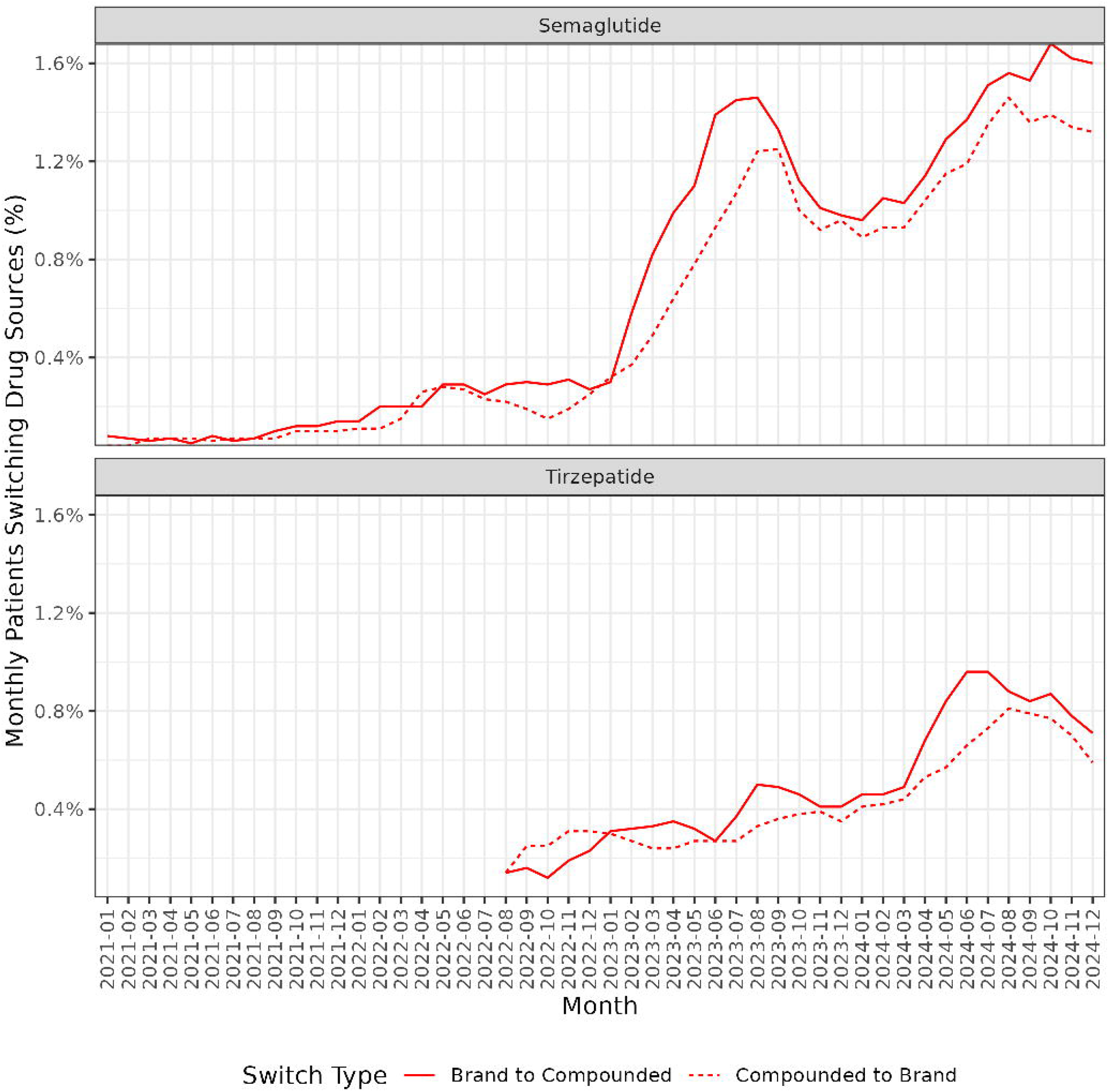

We observed significant differences between patients who used brand only versus those who used compounded drugs only (Table 1). Compounded-only patients were less likely to have a type 2 diabetes diagnosis – 13.8% versus 50.5% for brand-only patients – just as they were more likely to be female and non-Hispanic White. Users of compounded medications were more likely to live in states with Medicaid coverage of GLP-1 RAs for obesity – 92.2% of compounded-only users lived in these states, versus 84.1% of brand-only users – and lived in areas with significantly lower deprivation (20.2 for compounded-only users versus 30.2 for brand-only users). Notably, brand-only patients seemed to use the medications for a shorter duration than those who used compounded medications alone or switched between drug sources. While individuals who used only brand drugs had 7.8 (standard deviation [SD]: 10.5) months between their first and last documentation of medication use, compounded-only users had 9.9 (SD: 12.9) months.

**Table 1:** Characteristics of patients with documented semaglutide and/or tirzepatide use.

|  | **All**<br>**n=153,075** | **Both**<br>**n=6086** | **Brand Only**<br>**n=140,446** | **Compounded**<br>**Only**<br>**n=6543** |
| --- | --- | --- | --- | --- |
| *Female, N (%)\*\*\** | 98,027 (64.0%) | 4590 (75.4%) | 88660 (63.1%) | 4777 (73.0%) |
| *Non-Hispanic White, N (%)\*\*\** | 98,198 (64.2%) | 4081 (67.1%) | 89677 (63.9%) | 4440 (67.9%) |
| *Age, mean (SD)\*\*\** | 55.0 (13.7) | 51.4 (13.1) | 55.2 (13.5) | 53.4 (18.1) |
| *Type 2 diabetes diagnosis, N (%)\*\** | 73,128 (47.8%) | 1325 (21.8%) | 70903 (50.5%) | 900 (13.8%) |
| *Lives in state w/ Medicaid GLP-1 coverage for obesity, N (%)\*\** | 129,448 (84.6%) | 5316 (87.3%) | 118100 (84.1%) | 6032 (92.2%) |
| *Area deprivation index, mean (SD)\*\** | 29.6 (25.4) | 26.9 (21.6) | 30.2 (25.8) | 20.2 (13.9) |
| *Months between first and last documentation, mean (SD)\*\** | 8.09 (10.7) | 12.5 (11.3) | 7.81 (10.5) | 9.92 (12.9) |
| *A1c $\geq 6.5$, N (%)* | 29,457 (19.2%) | 596 (9.8%) | 28,428 (20.2%) | 433 (6.6%) |
| *A1C, mean (SD)\*\** | 7.01 (1.81) | 6.25 (1.44) | 7.09 (1.83) | 5.97 (1.17) |

Among switchers, differences were less pronounced (Table 2). Significantly fewer non-Hispanic White patients started with compounded drugs (63.8%) versus with brand drugs (68.2%). Those who initiated with compounded drugs were also more likely to have a diagnosis of type 2 diabetes (26.3% versus 21.9% for those starting with brand drugs). Similarly to the comparison of brand-only to compounded-only users, switchers who started with brand drugs lived in areas with higher deprivation levels (29.0) compared to those who started with compounded versions (22.2).

**Table 2:**
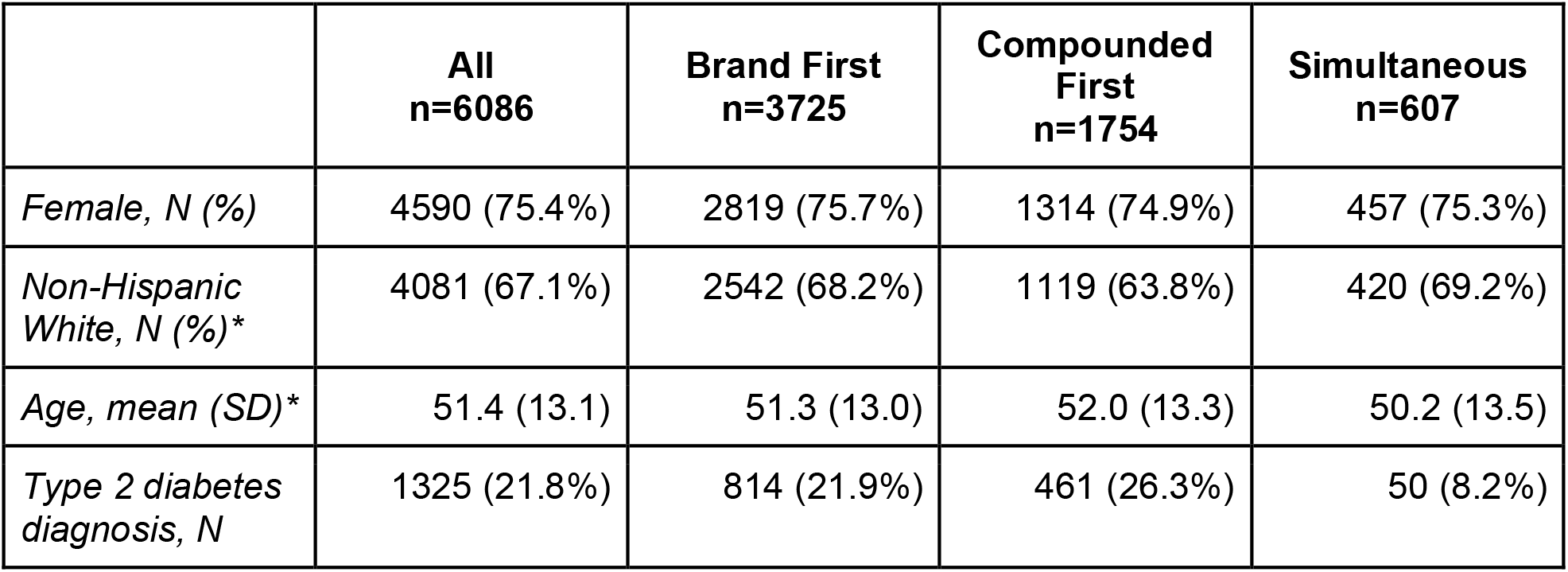
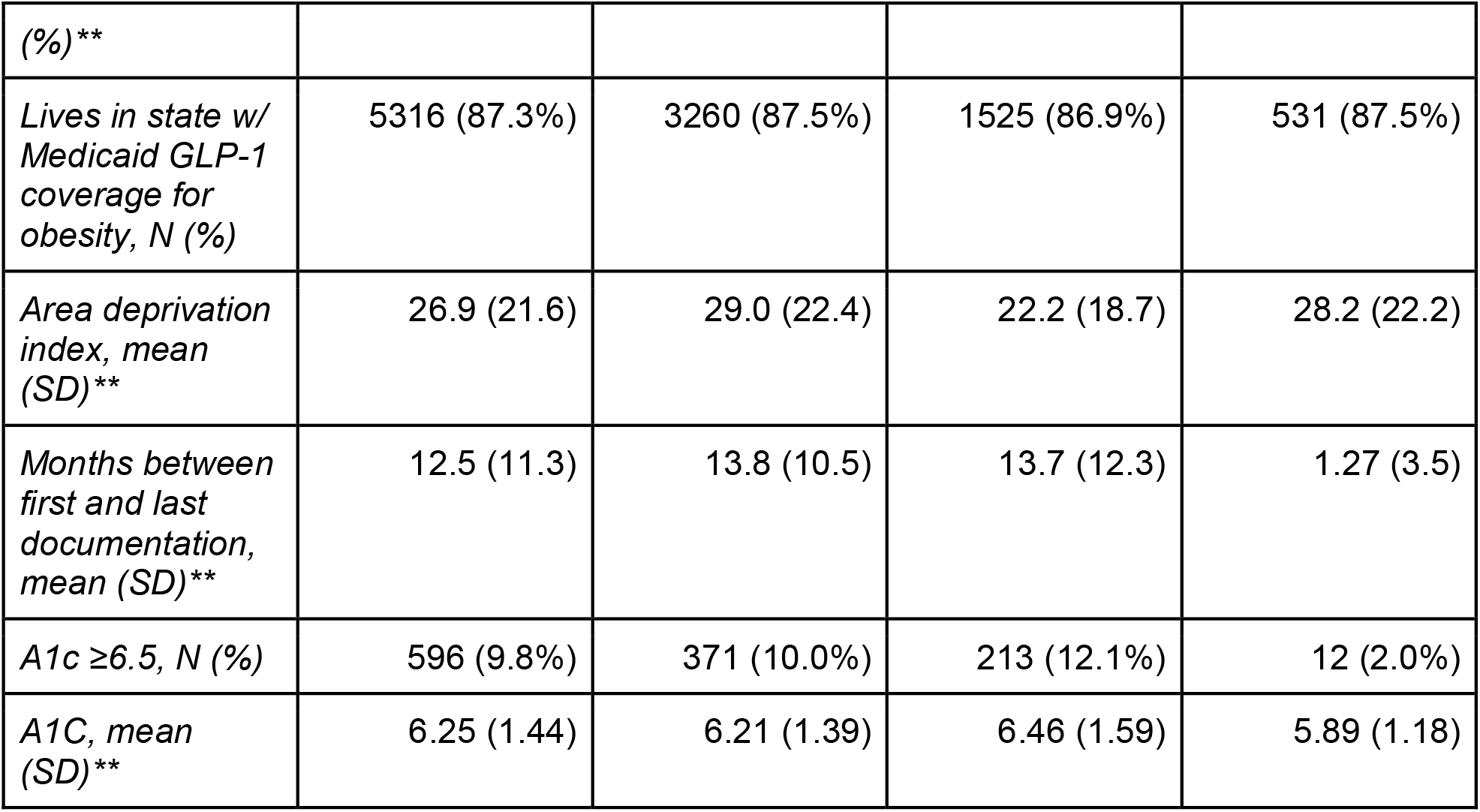
Characteristics of patients with documentation of both brand and compounded medications.

Our manual review of notes captured by our search strategy identified 5 false positives out of 500 sampled notes, indicating a positive predictive value of 99.0% (95% confidence interval: 97.7 to 99.6%).

## Discussion

In this study, we present novel data on documentation of compounded GLP-1 RAs in a large, nationwide cohort of primary care practices. We find that 8.3% of patients who used one of the two most popular GLP-1 RAs have documented use of compounded versions of these drugs. Relatively high rates of switching between drug sources may highlight widespread access difficulties stemming from supply and affordability issues. Furthermore, we observe significant differences in users of compounded drugs versus those who only use brand versions.

The need to search for documentation of compounded drugs in clinical notes highlights the inaccessibility of this data and the fragmented nature of the clinical ecosystem providing these drugs. Without interoperability between compounded drug providers’ records and clinical electronic health records, we cannot perform pharmacovigilance to ensure the safety and efficacy of compounded products, and clinicians may be unable to coordinate their patients’ medication use in alignment with evidence-based care. Notably, our finding that 8.3% of GLP-1 RA users have clinical documentation of compounded drugs is far short of the previously published estimate that 23% of users get prescriptions from someone other than their primary care provider or specialist. These higher estimates likely reflect patients obtaining compounded medication outside of traditional care; for example, through aesthetic or weight-loss service providers.

Our study has several limitations. First, our keyword search strategy for identification of compounded GLP-1 RA use prioritized high positive predictive value, likely at the expense of sensitivity, potentially underestimating true prevalence. Second, 5documentation may not reflect actual use. Since our records are not linked to pharmacy claims, we cannot confirm that patients filled any prescriptions. Third, without prescription end dates or refill data, we relied on documentation gaps to determine end dates for usage since we could not identify when prescriptions had been refilled. Finally, we are uncertain how our findings generalize across traditional primary care settings.

## Conclusion

In this novel national analysis of primary care documentation data, we used a high positive predictive value method for identifying compounded drug documentation from clinical notes within primary care, discovering compounded GLP-1 receptor agonist use in a meaningful minority of patients, at rates far lower than prior survey-based estimates. The discrepancy suggests significant patient receipt of these medications outside of coordinated or traditional primary care, and underscores the pressing need for clearer clinical guidance, regulatory oversight and interoperable data systems that can bridge gaps between whole-person primary care and newer channels of compounded medication access.

## Notes

### Competing Interest Statement

The authors have declared no competing interest.

### Funding Statement

The study did not receive any funding.

### Author Declarations

IRB of the American Academy of Family Physicians waived ethical approval for this work.

## References

1. Mattingly TJ II, Conti RM. Marketing and Safety Concerns for Compounded GLP-1 Receptor Agonists. JAMA Health Forum. 2025 Jan 17;6(1):e245015.

2. Chetty AK, Chillakanti M, Ramachandran R, Ross JS, Chen AS. Online Advertising of Compounded Glucagon-Like Peptide-1 Receptor Agonists. JAMA Health Forum. 2025 Jan 17;6(1):e245018.

3. Neumiller JJ, Bajaj M, Bannuru RR, McCoy RG, Pekas EJ, Segal AR, et al. Compounded GLP-1 and Dual GIP/GLP-1 Receptor Agonists: A Statement from the American Diabetes Association. Diabetes Care. 2024 Dec 2;48(2):177–81.

4. Montero A, Sparks G, Presiado M, Published LH. KFF Health Tracking Poll May 2024: The Public’s Use and Views of GLP-1 Drugs [Internet]. KFF. 2024 [cited 2025 Apr 10]. Available from: https://www.kff.org/health-costs/poll-finding/kff-health-tracking-poll-may-2024-the-publics-use-and-views-of-glp-1-drugs/

5. Vala A, Hao S, Chu I, Phillips RL, Rehkopf D. The American Family Cohort (v12.5). Stanford, CA: Redivis; 2023.

6. Wadden TA, Bailey TS, Billings LK, Davies M, Frias JP, Koroleva A, et al. Effect of Subcutaneous Semaglutide vs Placebo as an Adjunct to Intensive Behavioral Therapy on Body Weight in Adults With Overweight or Obesity: The STEP 3 Randomized Clinical Trial. JAMA. 2021 Apr 13;325(14):1403–13.

7. Rosenstock J, Wysham C, Frías JP, Kaneko S, Lee CJ, Fernández Landó L, et al. Efficacy and safety of a novel dual GIP and GLP-1 receptor agonist tirzepatide in patients with type 2 diabetes (SURPASS-1): a double-blind, randomised, phase 3 trial. The Lancet. 2021 Jul 10;398(10295):143–55.

8. Williams E, Rudowitz R, Published CB. Medicaid Coverage of and Spending on GLP-1s [Internet]. KFF. 2024 [cited 2025 Apr 15]. Available from: https://www.kff.org/medicaid/issue-brief/medicaid-coverage-of-and-spending-on-glp-1s/

9. Reproducible ADI (ReADI) | Socioeconomic Position Indices [Internet]. [cited 2025 Apr 17]. Available from: https://sepi.sites.stanford.edu/sep-indices/reproducible-adi-readi

